# Spinal mobilization characteristics: a scoping literature review of biomechanical parameters

**DOI:** 10.1101/2023.07.20.23292952

**Authors:** Lindsay M Gorrell, Luana Nyirö, Mégane Pasquier, Isabelle Pagé, Nicola R Heneghan, Petra Schweinhardt, Martin Descarreaux

## Abstract

**Background:** Spinal mobilization (SMob) is often included in the conservative management of spinal pain conditions as a recommended and effective treatment. While some studies quantify the biomechanical (kinetic) parameters of SMob, interpretation of findings is difficult due to poor reporting of methodological details. The aim of this study was to synthesise the literature describing biomechanical parameters of manually applied SMob.

**Methods:** This study is reported in accordance with the Preferred Reporting Items for Scoping Reviews (PRISMA-ScR) statement. Databases were searched from inception to October 2022: MEDLINE (Ovid), Embase, CINAHL, ICL, PEDro and Cochrane Library. Data were extracted and reported descriptively for the following domains: general study characteristics, number of and characteristics of individuals who delivered/received SMob, region treated, equipment used and biomechanical parameters of SMob.

**Results:** Of 7,607 records identified, 36 (0.5%) were included in the analysis. Of these, SMob was delivered to the cervical spine in 13 (36.1%), the thoracic spine in 3 (8.3%) and the lumbopelvic spine in 18 (50.0%) studies. In 2 (5.6%) studies, spinal region was not specified. For SMob applied to all spinal regions, biomechanical parameters were: peak force (0-128N); duration (10-120s); frequency (0.1-4.5Hz); and force amplitude (1-102N).

**Conclusions:** This study reports considerable variability of the biomechanical parameters of SMob. In studies reporting biomechanical parameters, SMob was most frequently delivered to the lumbar and cervical spine of humans and most commonly peak force was reported. Future studies should focus on the detailed reporting of biomechanical parameters to facilitate the investigation of clinical dose-response effects.

## INTRODUCTION

Musculoskeletal disorders, including low back and neck pain, affect most individuals during their lives (1–4). Such disorders are a prominent cause of disability globally (5) and can lead to decreased quality of life and psychological distress (6,7). The global prevalence of musculoskeletal disorders is increasing (8), as are their associated financial and societal costs (9–11). Individuals commonly seek care from various healthcare providers for the treatment of musculoskeletal disorders (12–14) and the use of evidence-based interventions is recommended (12,15–17). Clinical practice guidelines recommend the use of conservative treatments including spinal mobilization (SMob) and/or spinal manipulation (SM) for the treatment of musculoskeletal disorders (18–21). SMob is characterized by the manual application of oscillatory forces with low velocity and variable amplitude and frequency to an articulation (22). SMob can be further described in terms of the movement amplitude (or ’grade)’ as first described by Maitland (23). Specifically: Grade I involves a small-amplitude movement performed at the beginning of the range of motion (ROM); Grade II is a large-amplitude movement performed within the free range but not moving into any resistance or stiffness; Grade III is a large-amplitude movement performed up to the limit of the range; and Grade IV involves a small-amplitude movement performed at the limit of the range. It has been reported that Grades I and II SMob are often applied with the intention of pain reduction, while Grades III and IV are commonly used to increase ROM (24).

Transient neurophysiological effects in both the autonomic (e.g. changes in skin temperature and conductance (25,26)) and somatic (e.g. changes in muscle activity (26,27)) nervous systems have been reported in response to SMob. Additionally, beneficial clinical outcomes such as hypoalgesia (25,28–30) and increased ROM (27,31) have been linked to the intervention. However, it is yet to be established if physiological responses to manual therapy (i.e. SMob) are related to clinical outcomes (32). Therefore, the mechanisms underlying the beneficial clinical effects of SMob remain unclear (33,34) and without quantification of the intervention, it is difficult to determine which, if any, biomechanical parameters may influence patient outcomes (35).

To date, there have been two reviews of SMob biomechanical parameters (22,36) reporting on mean peak forces during SMob delivered in a posterior-anterior (PA) direction. In a 1997 review by Björnsdóttir and colleagues, force application was discussed in a single paragraph, with data reported for SMob delivered to: i) the L3 vertebra by 2 instructors (mean peak force: 33.3N); and ii) an unspecified thoracic level by 2 manual therapists using Grades I (means of the means for the 2 therapists: 134.75N) and IV (342.5N) (36). In a 2006 review, Snodgrass and colleagues evaluated the literature for consistency of force application by manual therapists during PA SMob (22). This review reported on mean peak forces in the PA direction for Grades I-IV SMob delivered to the spine (cervical:4; thoracic:3; and lumbar:7) and artificial devices (4). Both reviews highlighted a variability in nomenclature, definitions of biomechanical parameters and force delivery during SMob (22,36). Since the Snodgrass and colleagues review, there has been no further collation or synthesis of SMob biomechanical parameters. Therefore, the aim of this study was to synthesise the existing literature describing biomechanical (kinetic) parameters during the delivery of manually applied SMob.

## METHODOLOGY

This scoping literature review is reported in accordance with the Preferred Reporting Items for Scoping Reviews (PRISMA-ScR) statement (37). The protocol was developed by an experienced, international and interprofessional team and was prospectively registered at the Open Science Framework Registry (https://osf.io/3mqjs/). The original study design and subsequent search were conducted with the intention to capture information concerning the biomechanical parameters of both SMob and SM. Protocol deviations included that: i) due to the large quantity of data published on the topic, it was decided to report the biomechanical parameters of SMob and SM separately; and ii) studies reporting on SMob delivered to animals were excluded as it was unknown how biomechanically comparable SMob delivery would be to that delivered to humans. Due to the separate reporting of SMob and SM data in different manuscripts, several sections of the methods described here mirror those in the manuscript reporting on SM data (under peer-review).

### Eligibility criteria

Eligibility criteria were developed using the Sample, Phenomenon of Interest, Design, Evaluation, Research Type (SPIDER) search concept tool (38).

### Inclusion criteria

S – the sample population was humans (of any age) and non-humans (e.g. instrumented tool, manikin); PI – the phenomenon of interest was manually delivered SMob and/or SM, delivered by any regulated health professional (e.g. physiotherapist or chiropractor) or student enrolled at an accredited institution; D – observational study designs (e.g. case series studies, cohort and case-control studies); E – kinetic variables of the intervention (e.g. force-time profile); and R – original quantitative research data from studies utilizing SMob and/or SM as either the sole intervention or as a comparator.

### Exclusion criteria

Exclusion criteria were: i) SMob and/or SM delivered by a mechanical instrument or device; ii) all other therapeutic modalities; iii) manuscript not published in English, French or German; and iv) studies that had been retracted, were secondary analyses, trial registrations, protocols, clinical practice guidelines, commentaries, editorials, conference proceedings or single case studies.

### Search strategy

The search strategy was created by subject specific and methodological experts, with the assistance of an experienced medical and health sciences librarian. MEDLINE(Ovid), Embase, CINAHL, ICL, PEDro and Cochrane Library databases were searched from inception to 4 October 2022. The first author (LG) screened the reference lists of included studies to ensure that all relevant literature was captured. The following search terms and derivatives were adapted for each search engine: (spine, spinal, manipulation, mobilization or mobilisation, musculoskeletal, chiropractic, osteopathy, physiotherapy, naprapathy, force, motor skill, biomechanics, dosage, dose-response, education, performance, psychomotor, back, neck, spine, thoracic, lumbar, pelvic, cervical, sacral). Search strategies for all databases are provided in S1 Appendix.

### Study selection process

Records retrieved from the electronic searches were exported to the Rayyan© online platform (2022) (39) and duplicates were removed. Beginning with title and abstract review, groups of two authors (LG and LN; LG and IP; LG and MP) independently screened studies in a step-wise process. Full-texts of the remaining studies were then retrieved and screened independently by groups of two authors (LG and LN; LG and IP). Disagreements regarding study inclusion that could not be resolved by consensus were resolved by a third author (MD).

### Data extraction

Data were extracted from eligible studies by groups of two independent authors (LG and LN; LG and MP). These data included: i) general study characteristics (e.g. title, author, year and country of publication and type of study); ii) general study information (e.g. individual who delivered the intervention [e.g. clinician, student], professional qualification of individual delivering the intervention [e.g. physiotherapist, chiropractor], years of clinician experience/number of student hours, number of clinicians/students who delivered SMob or SM, recipient [e.g. human, manikin], number of recipients, whether the intervention was SMob [and grade of mobilization] or SM, the region treated [e.g. cervical, thoracic] and the measurement equipment used to record biomechanical parameters of the intervention); and iii) biomechanical parameters of SMob (e.g. peak force, SMob duration and frequency and force amplitude). Data reporting on SM is submitted for publication elsewhere (manuscript under peer review).

### Definitions

In this study, the following definitions were used:

- Peak force: the maximum applied force during a single SMob, reported as the mean of the force peaks that occurred during a specified period of the intervention.
- Duration: the time period of SMob delivery.
- Frequency: the rate of force oscillation during repeated applications.
- Force amplitude: the difference between the minimum and maximum forces applied during the intervention (i.e. the difference between a peak force and trough), reported as the mean of the force amplitudes that occurred during a specified period of SMob.
- Metrological details: descriptions of the suitability (e.g. accuracy, precision, sensitivity) of the measurement equipment to quantify the biomechanical parameters of SMob (40).

### Data synthesis

Descriptive statistics (mean, standard deviation and range) were used to report data. Any deviations from this (such as the use of 95% confidence intervals or the reporting of median and interquartile range) are explicitly indicated and reflect how the data were reported in the original studies. Microsoft Excel (Office 365, Microsoft Corporation, Redmond, USA) was used to calculate frequencies and proportions of trials reporting on each of the specified domains mentioned above.

In order to manage the substantial volume of data presented in this study, the following decisions were made regarding how to best report the data:: i) for studies reporting forces measured in 3-dimensions (3D) and including the resultant forces (i.e. the total forces applied), only the resultant forces are reported; ii) for studies measuring forces applied in 3D but not including the resultant forces, only the forces measured in the primary direction of the applied force are reported in the tables (e.g. for prone PA thoracic SMob, the vertical forces are reported). Regarding the reporting of metrological data, a consensus was reached by two authors (LG and MD) as to whether adequate information was provided. In cases where metrological details were discussed (e.g. it was stated that measurement equipment accuracy was good) but it was not clear if this statement was based on data (or what data), this was recorded as metrological details were not provided. No assessment of study quality was performed.

## RESULTS

The electronic searches returned 7,607 records, with 3,981 unique records remaining after de- duplication (n=3,626) (Figure 1). Following title/abstract screening, 247 full-texts were screened. Of these, 146 reports were excluded (e.g. did not report biomechanical parameters: 56), leaving 101 included studies. Of these, 36 reported on SMob and were included in the analysis. A list of these studies is provided in S2 Appendix and the reference number cited in the tables refers to this list.

**Figure 1:**
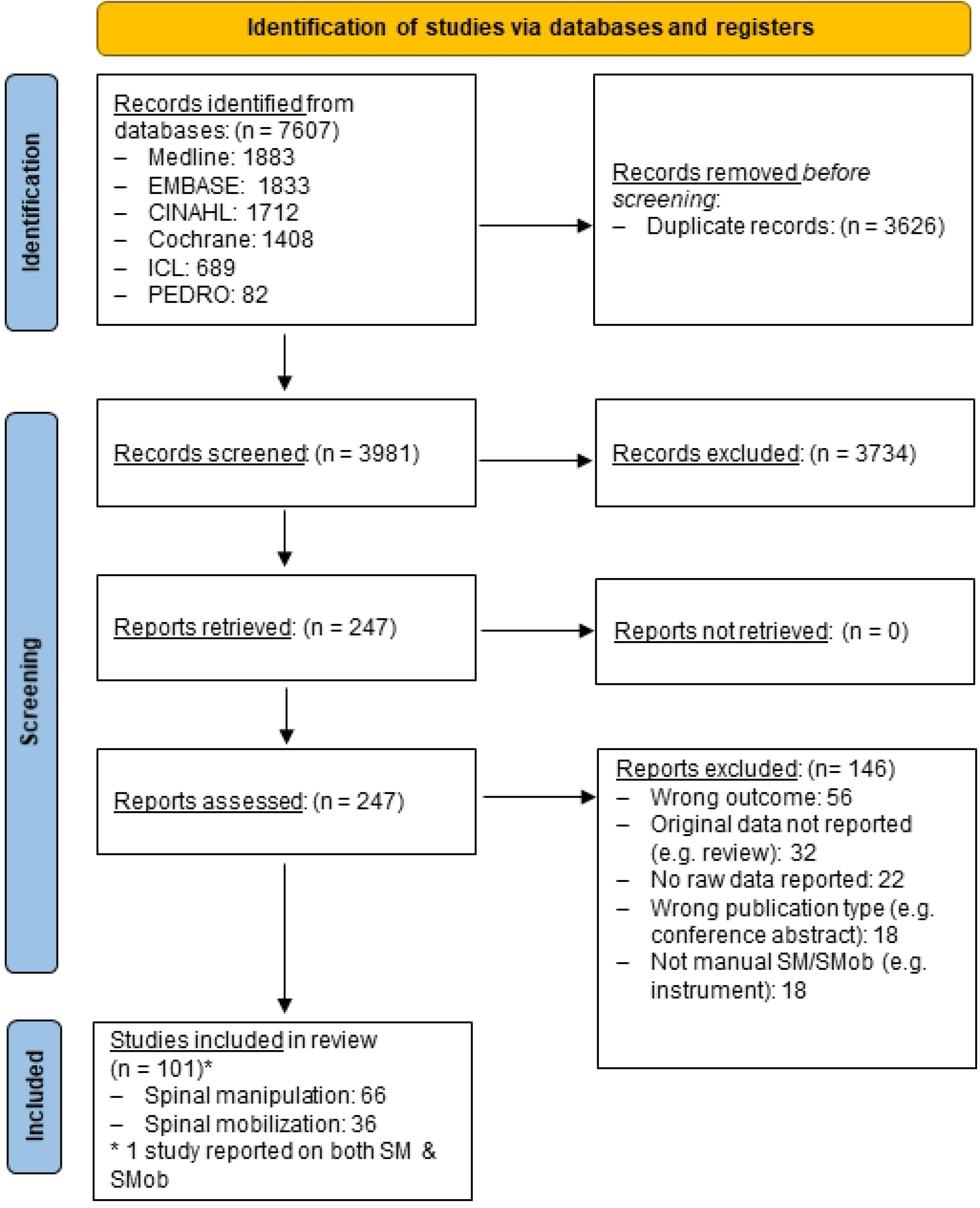
PRISMA flow-chart.

Of the 36 included studies, most were published in the 10-year period from 2003 to 2012 (n=13, 36.1%) in Australia (n=13, 36.1%) (Table 1). Typically, the study design was cross-sectional (n=26, 72.2%), with SMob delivered by a clinician only (i.e. no students were involved) (n=24, 66.7%) whose profession was a physiotherapist (n=27, 75.0%). In the 31 (86.1%) studies in which SMob was delivered by clinicians, clinical experience was unclear in 14 (45.2%) studies and when it was reported, clinicians with more than 5 years’ experience most commonly delivered SMob (n=10, 32.3%). When SMob was delivered by a student (n=12, 33.3%), the number of SMob training hours was not reported in any (n=12, 100.0%) study. Most frequently, the number of individuals (i.e. clinicians and/or students) delivering SMob was between 1 and 49 (n=29, 80.6%), with 12 (33.3%) of these studies involving only 1 to 2 individuals delivering SMob. SMob was delivered to adults (18 to 65 years) in 20 (55.6%) studies, with the demographics of the cohort to which SMob was delivered not reported in 8 (22.2%) studies. The number of individuals receiving SMob was reported as between 1 and 49 in 28 (77.8%) studies, with only 1 to 2 individuals receiving SMob in 10 (27.8%) studies. SMob was most commonly delivered to the lumbopelvic spine (n=18, 50.0%) and the cervical spine (n=13, 36.1%), and the SMob ’technique’ was reported in all but one study (n=35, 97.2%). Biomechanical parameters were measured at the patient-table interface in 16 (44.4%) studies, another interface (e.g. thumbnail of the clinician) in 6 (16.7%) studies, the clinician-patient interface in 5 (13.9) studies and the clinician-ground interface in 4 (11.1%) studies. Metrological data of the measurement equipment were reported in 27 (75.0%) studies. Regarding biomechanical parameters, the following were reported: peak force in 35 (97.2%); SMob duration in 12 (33.3%); SMob frequency in 16 (44.4%); and amplitude of force in 11 (30.6%) studies.

**Table 1:** Overall summary of studies reporting on the biomechanical parameters of spinal mobilization (SMob) (n=36).

|  | n, (%) |  | n, (%) |
| --- | --- | --- | --- |
| Year, n=36 |  | Individual who received SMob, n=36 |  |
| 2013 to 2022 | 12 (33.3) | Adult (18-65yr) | 20 (55.6) |
| 2003 to 2012 | 13 (36.1) | Geriatric (>65yr) | 1 (2.8) |
| 1993 to 2002 | 10 (27.8) | Instrumented tool/force plate | 4 (11.1) |
| Before 1993 | 1 (2.8) | Manikin | 1 (2.8) |
| Country, n=36 |  | Mixed | 2 (5.6) |
| Australia | 13 (36.1) | Unclear | 8 (22.2) |
| Canada | 6 (16.7) | Number of individuals receiving SMob, n=36 |  |
| England | 6 (16.7) | 1 or 2 | 10 (27.8) |
| Ireland | 1 (2.8) | 0 to 49 | 28 (77.8) |
| Malaysia | 1 (2.8) | 50 to 99 | 5 (13.9) |
| South Africa | 1 (2.8) | Not reported | 3 (8.3) |
| Unclear | 1 (2.8) | Region SMob delivered to, n=36 |  |
| USA | 7 (19.4) | Cervical | 13 (36.1) |
| Study type, n=36 |  | Thoracic | 3 (8.3) |
| Cross-sectional | 26 (72.2) | Lumbopelvic | 18 (50.0) |
| Prospective | 10 (27.8) | Other | 2 (5.6) |
| Individual who delivered SMob, n=36 |  | Technique reported, n=36 |  |
| Practitioner | 24 (66.7) | Yes | 35 (97.2) |
| Student | 5 (13.9) | No | 1 (2.8) |
| Both | 5 (13.9) | Measurement interface, n=36 |  |
| Unclear | 2 (5.6) | Patient-table | 16 (44.4) |
| Profession, n=36 |  | Clinician-patient | 5 (13.9) |
| Physiotherapist | 27 (75.0) | Clinician-ground | 4 (11.1) |
| Chiropractor | 5 (13.9) | Table-ground | 3 (8.3) |
| Unclear | 4 (11.1) | Both clinician-patient & patient-table | 1 (2.8) |
| Experience (clinician) n=31 |  | Other | 6 (16.7) |
| >5yr | 10 (32.3) | Unclear | 3 (8.3) |
| Mixed | 7 (22.6) | Metrological data reported, n=36 |  |
| Unclear | 14 (45.2) | Reported | 27 (75.0) |
| Hours of training (student) n=12 |  | Not reported | 9 (25.0) |
| Reported | 0 (0.0) | Peak force, n=36 |  |
| Not reported | 12 (100.0) | Reported | 35 (97.2) |
| Number of individuals delivering SMob, n=36 |  | Not reported | 1 (2.8) |
| 1 or 2 | 12 (33.3) | Duration of mobilization, n=36 |  |
| 1 to 49 | 29 (80.6) | Reported | 12 (33.3) |
| 50 to 99 | 2 (5.6) | Not reported | 24 (66.7) |
| 100 to 149 | 2 (5.6) | Frequency of mobilization, n=36 |  |
| >150 | 1 (2.8) | Reported | 16 (44.4) |
| Not reported | 2 (5.6) | Not reported | 20 (55.6) |
|  |  | Amplitude of force, n=36 |  |
|  |  | Reported | 11 (30.6) |
|  |  | Not reported | 25 (69.4) |

Abbreviations: n: number of studies, SMob: spinal mobilization, y: year, >: greater than, <: less than.

### Cervical spine

Of the 11 (84.6%) studies that reported on SMob delivered to the cervical spine of humans, the following biomechanical parameters were reported: i) peak force: 0-128N; ii) duration: 60s; iii) frequency: 0.28-2.4Hz; and iv) force amplitude: 14.4-52.5N (Tables 2 & 3; S3 Appendix, Table A). Of the 2 (15.4%) studies that reported on SMob delivered to non-humans (i.e. human analogue manikin:1; instrumented tool:1) peak force 42-181N was reported.

**Table 2:** Summary of studies reporting on the biomechanical parameters of spinal mobilization (SMob) delivered to the cervical spine of humans (n=11) and non-humans (i.e. human analogue manikins, instrumented tools) (n=2).

| Author/s<br>Year, Country | SMob delivery<br>Profession (n) | Experience | Recipient/s (n) | Location/s | Technique/s<br>Grade/s | Interface/s | Equipment | Metrological<br>data |
| --- | --- | --- | --- | --- | --- | --- | --- | --- |
| **Humans** |  |  |  |  |  |  |  |  |
| Conradie et al<br>2004, South Africa<sup>7</sup> | Clin<br>Physio (16) | NR | Adult (1) | C6 | PA<br>I | Clin-pat | Pressure sensor | No |
| Snodgrass et al<br>2006, Australia<sup>28</sup> | Clin<br>Physio (10) | Mixed | Adult (1) | C2/C7 | PA<br>I-IV | Pat-table | Load cells | No |
| Snodgrass et al<br>2009, Australia<sup>29</sup> | Clin<br>Physio (116) | >5y | Adult (35) | C2/C7 | PA<br>I-IV | Pat-table | Load cells | Yes |
| Snodgrass et al<br>2010, Australia<sup>30,31</sup> | Clin & Stud<br>Physio (336) | Clin: NR<br>Stud: NR (no<br>clin exper) | Adult (67) | C2/C7 | PA<br>I-IV | Pat-table | Load cells | Yes |
| Snodgrass et al<br>2010, Australia<sup>31</sup> | Stud<br>Physio (120) | NR (2-4 y) | Adult (32) | C2/C7 | PA<br>I-IV | Pat-table | Load cells | Yes |
| Gudavalli et al<br>2013, USA<sup>13</sup> | Clin<br>Chiro (4) | NR | Adult (9) | C5/C6 | C distract<br>NA | Pat-table | Force plate | No |
| Snodgrass et al<br>2014, Australia<sup>33</sup> | Clin<br>Physio (1) | >5y | Adult (64) | MP (C3-7) | PA<br>III | Pat-table | Load cells | No |
| Gudavalli et al<br>2015, USA<sup>15</sup> | Clin<br>Chiro (NR) | NR | Adult (45) | Occi/C5 | C distract<br>NA | Pat-table | Force plate | No |
| Gudavalli et al<br>2015, USA<sup>16</sup> | Clin<br>Chiro (2) | >5y | Adult (48) | Occi/C5 | C distract<br>NA | Pat-table | Force plate | No |
| Kope et al<br>2018, Canada<sup>19</sup> | Clin<br>Physio (5) | >5y | NR (NR) | C5/C6 | PA<br>III | Clin thumbnail | Strain gauge | No |
| Chia et al<br>2021, Malaysia<sup>4</sup> | Clin<br>Physio (1) | NR | NR (30) | C6 | PA<br>I-IV | NR | Pressure sensor | No |
| **Non-humans** |  |  |  |  |  |  |  |  |
| Buckingham et al<br>2007, Australia<sup>3</sup> | Clin & Stud<br>Physio (36) | Clin: Mixed<br>Stud: NR (4<sup>th</sup> y) | Manikin (1) | C6 | PA<br>NR | In manikin | Strain gauge | No |
| Walsh et al<br>2011, Ireland<sup>35</sup> | Stud<br>Physio (40) | NR (final y) | Instrumented<br>tool (1) | C7 | PA<br>III | Instrumented tool | Pinch-grip<br>analyser | No |

Abbreviations: C: cervical, Chiro: chiropractor, Clin: clinician, distract: distraction, exper: experience, Mixed: experience of clinicians both > and < 5 years, MP: most painful, (n): number of participants, NA: not applicable, NR: not reported, Occi: occiput, PA: posterior-anterior, Pat: patient, Physio: physiotherapist, SMob: spinal mobilization, Stud: students, y: years, >: greater than.

**Table 3:** Summary of biomechanical parameters reported by region for studies reporting on spinal mobilization (SMob) (n=36).

|  | Location of measurement<br>n (%) | Metrologic data<br>reported<br>n (%) | Peak force reported n<br>(%)<br>[range (N)] | Duration reported<br>n (%)<br>[range (s)] | Frequency reported n<br>(%)<br>[range (Hz)] | Force amplitude<br>reported n (%)<br>[range (N)] |
| --- | --- | --- | --- | --- | --- | --- |
| Humans<br>(n=11) | Patient-table: 8 (72.7)<br>Clinician-patient: 1 (9.1)<br>NR: 1 (9.1)<br>Other: 1 (9.1) | 3 (27.3) | 11 (100.0)<br>[0-128] | 1 (9.1)<br>60 | 6 (54.5)<br>[0.3-2.4] | 4 (36.4)<br>[14.4-52.5] |
| Non-humans<br>(n=2) | Within device: 1 (50.0)<br>Clinician-device: 1 (50.0) | 0 (0.0) | 2 (100.0)<br>[42-181] | 0 (0.0)<br>[NA] | 0 (0.0)<br>[NA] | 0 (0.0)<br>[NA] |
| Humans<br>(n=2) | Clinician-patient: 1 (50.0)<br>Clinician-patient &<br>patient-table: 1 (50.0) | 1 (50.0) | 1 (50.0)<br>[297-323] | 1 (50.0)<br>3x60 | 1 (50.0)<br>[28-32<sup>Δ</sup>] | 0 (0.0)<br>[NA] |
| Non-humans<br>(n=1) | Clinician-device<br>& table-ground: 1 (100.0) | 0 (0.0) | 1 (100.0)<br>[106-223] | 0 (0.0)<br>[NA] | 1 (100.0)<br>0.5 | 0 (0.0)<br>[NA] |
| Humans<br>(n=17) | Patient-table: 7 (41.2)<br>Clinician- ground: 4<br>(23.5)<br>Table-ground: 3 (17.6)<br>Clinician-patient: 2 (11.8)<br>NR: 1 (5.9) | 3 (17.6) | 16 (94.1)<br>[3-430] | 8 (47.1)<br>[10-120] | 7 (41.2)<br>[0.1-4.5] | 7 (41.2)<br>[1-102] |
| Non-humans<br>(n=1) | Device-table: 1 (100.0) | 0 (0.0) | 1 (100.0)<br>[36-119] | 1 (100.0)<br>30 | 0 (0.0)<br>[NA] | 0 (0.0)<br>[NA] |
| Non-humans<br>(n=2) | Within device: 1 (50.0)<br>Clinician-device &<br>Clinician-ground: 1<br>(50.0) | 0 (0.0) | 2 (100.0)<br>[2-361] | 1 (50.0)<br>20 | 1 (50.0)<br>[28-32<sup>Δ</sup>] | NR |

Abbreviations: Hz: Hertz, N: Newtons, n: number of studies, NA: not applicable, NR: not reported, Other: clinician thumb nailbed, s: seconds, SMob: spinal mobilization, Δ: data are reported as cycles/minute.

### Thoracic spine

Of the 2 (66.7%) studies that reported on SMob delivered to the thoracic spine of humans, the following biomechanical parameters were reported: i) peak force: 297-323N; ii) duration: 3×60s; and iii) frequency: 0.47-0.53Hz (Tables 3 & 4; S3 Appendix, Table B). In the one (33.3%) study that reported on SMob delivered to 12 T5-8 sections of human cadavers: i) peak force: 106-223N; and ii) frequency: 0.5Hz were reported.

**Table 4:** Summary of studies reporting on the biomechanical parameters of spinal mobilization (SMob) delivered to the thoracic spine of humans (n=2) and non-humans (i.e. partial cadaveric sections) (n=1).

| Author/s<br>Year, Country | SMob delivery<br>Profession (n) | Experience | Recipient/s (n) | Location/s | Technique/s<br>Grade/s | Interface/s | Equipment | Metrological<br>data |
| --- | --- | --- | --- | --- | --- | --- | --- | --- |
| **Humans** |  |  |  |  |  |  |  |  |
| Zegarrra-Parodi et al<br>2016, NR<sup>36</sup> | NR<br>NR (1) | NR | Adult (32) | T1 | Lateral glide via lamina<br>NR (5/40/80% of PPT) | Clin-pat | Pressure sensor | No |
| Funabashi et al<br>2021, Canada<sup>10</sup> | Clin<br>Chiro (1) | >5y | Geriatric (18) | T1-12 | Clin choice<br>4 | Clin-pat<br>Pat-table | Load cells<br>Force plate | Yes |
| **Non-humans** |  |  |  |  |  |  |  |  |
| Sran et al<br>2004, Canada<sup>34</sup> | Clin<br>Physio (2) | NR | Cadaveric<br>sections T5-8<br>(12) | T6 | PA<br>NR | Clin-pat<br>Table-floor | Pressure sensor | No |

Abbreviations: Chiro: chiropractor, Clin: clinician, (n): number of participants, NR: not reported, PA: posterior-anterior, Pat: patient, Physio: physiotherapist, PPT: pressure pain threshold, SMob: spinal mobilization, T: thoracic, y: years, >: greater than.

### Lumbopelvic spine

Of the 17 (94.4%) studies that reported on SMob delivered to the lumbar spine of humans, the following biomechanical parameters were reported: i) peak force: 3-430N; ii) duration: 10-120s; iii) frequency: 0-5Hz; and iv) force amplitude: 1-102N (Tables 3 & 5; S3 Appendix, Table C). In the one (33.3%) study that reported on SMob delivered to an instrumented tool: i) peak force: 36-119N; and ii) duration: 30s were reported.

**Table 5:**
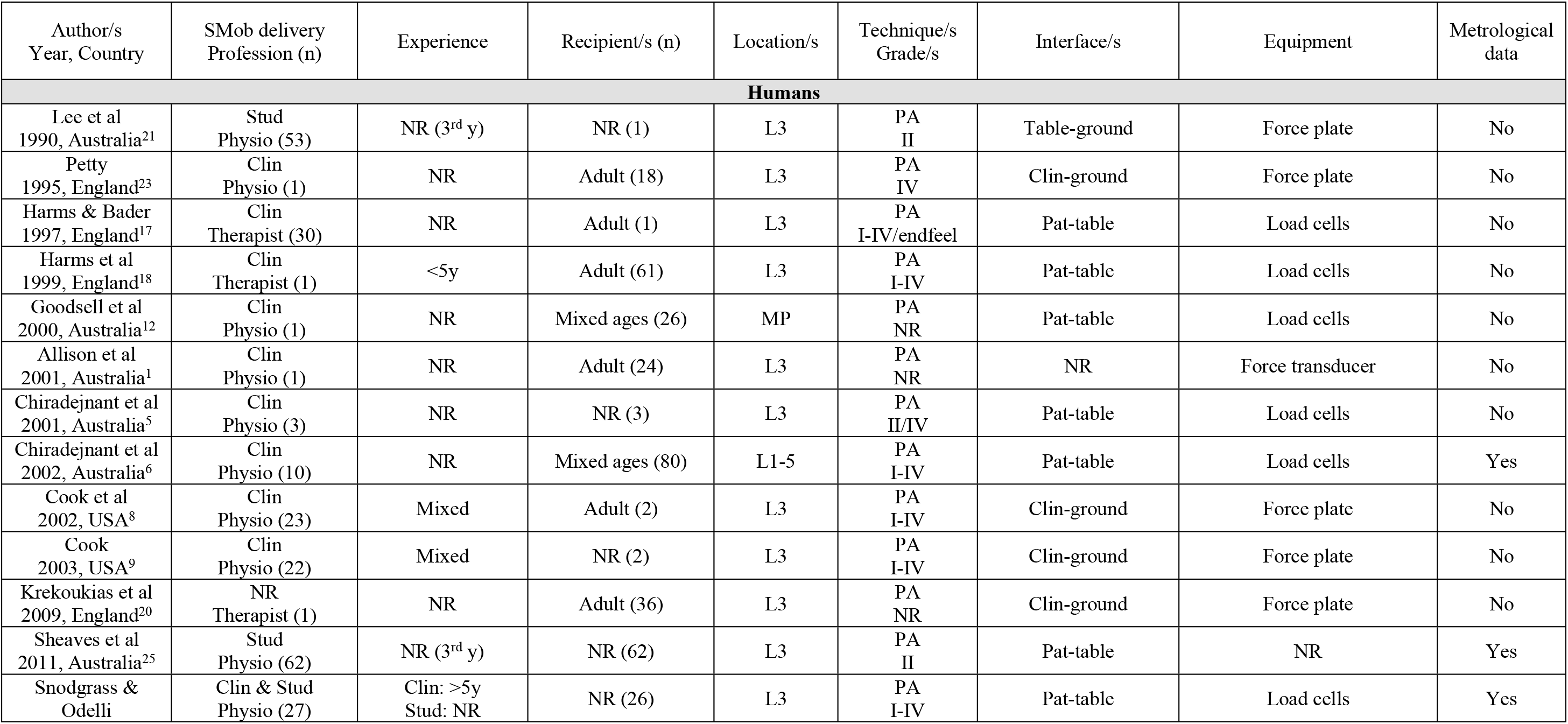
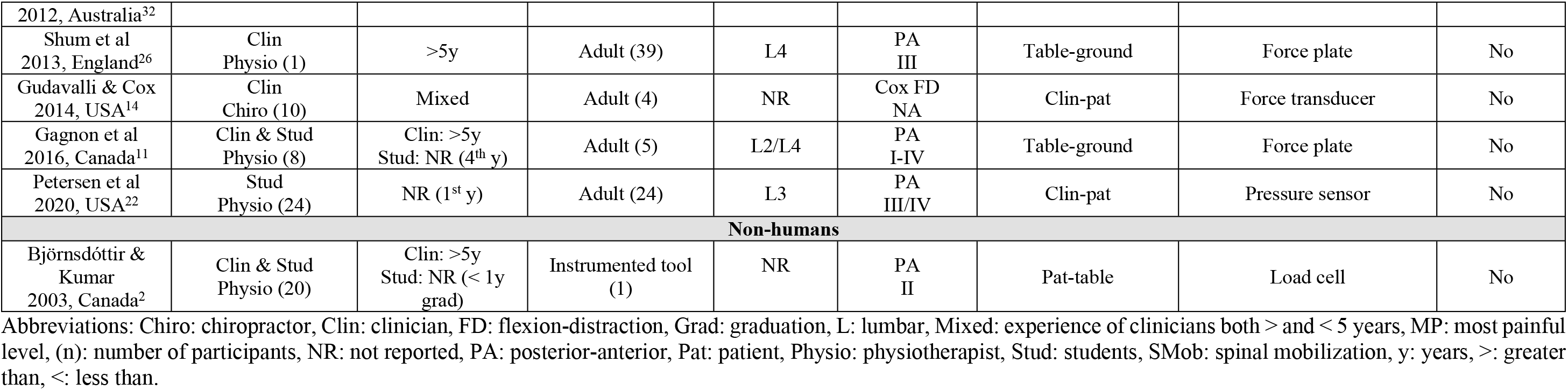
Summary of studies reporting on the biomechanical parameters of spinal mobilization (SMob) delivered to the lumbopelvic spine of humans (n=17) and non-humans (i.e. instrumented tools) (n=1).

| Author/s<br>Year, Country | SMob delivery<br>Profession (n) | Experience | Recipient/s (n) | Location/s | Technique/s<br>Grade/s | Interface/s | Equipment | Metrological<br>data |
| --- | --- | --- | --- | --- | --- | --- | --- | --- |
| **Humans** |  |  |  |  |  |  |  |  |
| Lee et al<br>1990, Australia<sup>21</sup> | Stud<br>Physio (53) | NR (3<sup>rd</sup> y) | NR (1) | L3 | PA<br>II | Table-ground | Force plate | No |
| Petty<br>1995, England<sup>23</sup> | Clin<br>Physio (1) | NR | Adult (18) | L3 | PA<br>IV | Clin-ground | Force plate | No |
| Harms & Bader<br>1997, England<sup>17</sup> | Clin<br>Therapist (30) | NR | Adult (1) | L3 | PA<br>I-IV/endfeel | Pat-table | Load cells | No |
| Harms et al<br>1999, England<sup>18</sup> | Clin<br>Therapist (1) | <5y | Adult (61) | L3 | PA<br>I-IV | Pat-table | Load cells | No |
| Goodsell et al<br>2000, Australia<sup>12</sup> | Clin<br>Physio (1) | NR | Mixed ages (26) | MP | PA<br>NR | Pat-table | Load cells | No |
| Allison et al<br>2001, Australia<sup>1</sup> | Clin<br>Physio (1) | NR | Adult (24) | L3 | PA<br>NR | NR | Force transducer | No |
| Chiradejnant et al<br>2001, Australia<sup>5</sup> | Clin<br>Physio (3) | NR | NR (3) | L3 | PA<br>II/IV | Pat-table | Load cells | No |
| Chiradejnant et al<br>2002, Australia<sup>6</sup> | Clin<br>Physio (10) | NR | Mixed ages (80) | L1-5 | PA<br>I-IV | Pat-table | Load cells | Yes |
| Cook et al<br>2002, USA<sup>8</sup> | Clin<br>Physio (23) | Mixed | Adult (2) | L3 | PA<br>I-IV | Clin-ground | Force plate | No |
| Cook<br>2003, USA<sup>9</sup> | Clin<br>Physio (22) | Mixed | NR (2) | L3 | PA<br>I-IV | Clin-ground | Force plate | No |
| Krekoukias et al<br>2009, England<sup>20</sup> | NR<br>Therapist (1) | NR | Adult (36) | L3 | PA<br>NR | Clin-ground | Force plate | No |
| Sheaves et al<br>2011, Australia<sup>25</sup> | Stud<br>Physio (62) | NR (3<sup>rd</sup> y) | NR (62) | L3 | PA<br>II | Pat-table | NR | Yes |
| Snodgrass &<br>Odelli | Clin & Stud<br>Physio (27) | Clin: >5y<br>Stud: NR | NR (26) | L3 | PA<br>I-IV | Pat-table | Load cells | Yes |
| 2012, Australia<sup>32</sup> |  |  |  |  |  |  |  |  |
| Shum et al<br>2013, England<sup>26</sup> | Clin<br>Physio (1) | >5y | Adult (39) | L4 | PA<br>III | Table-ground | Force plate | No |
| Gudavalli & Cox<br>2014, USA<sup>14</sup> | Clin<br>Chiro (10) | Mixed | Adult (4) | NR | Cox FD<br>NA | Clin-pat | Force transducer | No |
| Gagnon et al<br>2016, Canada<sup>11</sup> | Clin & Stud<br>Physio (8) | Clin: >5y<br>Stud: NR (4<sup>th</sup> y) | Adult (5) | L2/L4 | PA<br>I-IV | Table-ground | Force plate | No |
| Petersen et al<br>2020, USA<sup>22</sup> | Stud<br>Physio (24) | NR (1<sup>st</sup> y) | Adult (24) | L3 | PA<br>III/IV | Clin-pat | Pressure sensor | No |
| **Non-humans** |  |  |  |  |  |  |  |  |
| Björnsdóttir &<br>Kumar<br>2003, Canada<sup>2</sup> | Clin & Stud<br>Physio (20) | Clin: >5y<br>Stud: NR (< 1y<br>grad) | Instrumented tool<br>(1) | NR | PA<br>II | Pat-table | Load cell | No |

Abbreviations: Chiro: chiropractor, Clin: clinician, FD: flexion-distraction, Grad: graduation, L: lumbar, Mixed: experience of clinicians both > and < 5 years, MP: most painful level, (n): number of participants, NR: not reported, PA: posterior-anterior, Pat: patient, Physio: physiotherapist, Stud: students, SMob: spinal mobilization, y: years, >: greater than, <: less than.

### No region specified

Of the 2 (5.6%) studies that reported on SMob delivered to an unspecified region, the following biomechanical parameters were reported: i) peak force: 2-361N; ii) duration: 20s; and iii) frequency: 28-32 cycles/min (Tables 3 & 6; S3 Appendix, Table D).

**Table 6:** Summary of studies reporting on the biomechanical parameters of spinal mobilization (SMob) delivered to non-humans (i.e. instrumented tools) with no region specified (n=2).

| Author/s<br>Year, Country | SMob delivery<br>Profession (n) | Experience | Recipient/s (n) | Location/s | Technique/s<br>Grade/s | Interface/s | Equipment | Metrological data |
| --- | --- | --- | --- | --- | --- | --- | --- | --- |
| **Non-humans** |  |  |  |  |  |  |  |  |
| Simmonds et al<br>1995, Canada<sup>27</sup> | Clin<br>Physio (10) | >5y | Instrumented tool (1) | NA | PA<br>I-V | In tool | Pinch-grip analyser | No |
| Petty & Messenger<br>1996, England<sup>24</sup> | Clin<br>Physio (1) | NR | Instrumented tool (1) | NA | PA<br>NR | Clin-tool<br>Clin-ground | Pinch-grip analyser<br>Force platform | No |

Abbreviations: Clin: clinician, (n): number of participants, NA: not applicable, NR: not reported, PA: posterior-anterior, Physio: physiotherapist, SMob: spinal mobilization, y: years, >: greater than.

## DISCUSSION

This scoping review comprehensively synthesizes the existing evidence describing biomechanical parameters during the delivery of manually applied SMob, underscoring the substantial variability observed in these parameters. This finding is consistent with the results of two previous reviews reporting on SMob peak forces delivered in a PA direction (22,36), despite the current study having a larger scope of reporting (i.e. duration, frequency and force amplitude in addition to peak forces). The observed heterogeneity in the reported biomechanical parameters of SMob is likely attributable to several factors, such as differences in the: i) ’technique’ used (e.g. PA vs oscillatory distractive techniques); ii) measurement equipment used (e.g. force plate vs. pressure pad); iii) location of biomechanical parameter measurement (e.g. clinician-patient vs. patient-table); iv) individuals/equipment to which SMob was applied (e.g. body mass index (BMI), equipment materials); and v) individuals who delivered SMob (e.g. experience). However, without detailed descriptions of each of these domains, it is not possible to know if the identified biomechanical parameter variability is related to methodological and/or reporting differences, inherent differences in SMob application or, some other factor (41). Additionally, with the continued increase in the reporting on SMob biomechanical parameter data, it is imperative that detailed descriptions are reported in manuscripts to allow readers the opportunity to assess for themselves possible reasons for differences in the data.

### Detailed reporting of the intervention is necessary

It is unknown if meaningful clinical differences between different types of SMob ’technique’ (e.g. PA vs. oscillatory distraction) and/or grades of SMob exist (35). To assess this in future studies, there should be a detailed description of the applied SMob, including an explicit explanation of how the grade of SMob was defined. In the current review, detailed descriptions of SMob were not consistently provided. Specifically, there was large variation in the detail of SMob reporting and in many studies, replication of the intervention would be impossible based on the (lack of) details provided. Despite the existence of established reporting guidelines for health-related interventions (41), this lack of detail is not unique to SMob, existing also for interventions including SM (SM manuscript under peer-review) and dry needling (48). It is recommended that a specific guideline for the standardized reporting of SMob, similar to that for the standardized reporting of SM (49), is developed to improve the reporting of SMob interventions. Indeed the (development and) use of such a guideline would go some way to improving the generally poor level of manual therapy trials reporting (50).

### Factors possibly responsible for reported heterogeneity of SMob biomechanical parameters

It is not yet fully understood why forces measured at the clinician-patient and patient-table interfaces differ (42,43). However, as differences do exist, it is important for authors to report where and how biomechanical parameters were measured as results from studies measuring forces at different interfaces are not directly comparable. Furthermore, the terminology used should enable replication of study methodologies in future investigations and would be facilitated by the detailed reporting of biomechanical parameter definitions and calculations (e.g. was the peak force measured directly by the equipment or, was it mathematically estimated?). Without such details, it is impossible to compare biomechanical parameters between studies. Furthermore, it has been reported that demographics (e.g. sex, height and BMI) influence the application of manual therapy treatments (i.e. SMob and SM) (44–47). Specifically, these interventions are delivered more forcefully to males with a higher BMI. In summary, the interface of measurement, equipment used, terminology and SMob recipients should be systematically reported in detail to allow for both replication in future studies and reader judgement of the clinical relevance of reported results.

### Do biomechanical parameters matter clinically?

It has been reported that biomechanical parameters of SMob (e.g. peak force and force amplitude) differ between students and experienced clinicians (45). Specifically, students apply less force, more slowly. Similar differences are also reported in the SM literature (51), further suggesting that the detailed reporting of the clinical experience of the individual delivering the intervention is necessary. Furthermore, Gorgos and colleagues reported on the reliability of inter-clinician and intra-clinician forces applied during joint mobilisation in a systematic review (52). The authors concluded that while there is variability in the application of force between different clinicians, individual clinicians apply forces consistently. Despite such between-clinician differences in SMob force application, the literature shows that recipients experience beneficial clinical outcomes from various forms of manual therapy (including SMob) despite considerable differences in the biomechanical parameters of the applied interventions (e.g. low velocity, variable amplitude SMob vs. high velocity, low amplitude SM) (35,53,54). Additionally, some authors suggest that there is a threshold of ’dosage’ (in terms of biomechanical parameters such as force and/or rate of force application), rather than an optimal intervention approach (i.e. SMob vs SM), required to elicit beneficial clinical outcomes (55,56). However, to our best knowledge, this subject has not been systematically investigated and no reference standards (i.e. ranges of biomechanical parameters) for SMob application have been published (52). Furthermore, the lack of detailed description of biomechanical parameters limits the generalizability of results reported by both individual studies, and their subsequent syntheses, to clinical practice as it remains unclear exactly what ’dosage’ was applied (48). By exhaustively collating the existing literature, the current review provides a first step towards the development of such reference standards. However, the systematic biomechanical quantification of SMob is required to first establish if ’dosage’ is related to physiological responses (e.g. changes in the autonomic and somatic nervous systems) and/or clinical outcomes (e.g. hypoalgesia).

### Strengths and limitations

This review is the first to synthesise SMob biomechanical parameter data beyond peak force (including also duration, frequency and force amplitude) and includes 21 additional studies since the publication of the most recent 2006 review (22). The review was conducted by an international and interprofessional team and reported according to the (PRISMA-ScR) statement (37). The study provides a first step towards the systematic and detailed reporting of SMob interventions, which is necessary to investigate the relationship between the application of SMob and its’ observed clinical outcomes.

It is possible that there was unintentional exclusion of studies reporting on the parameters of interest. However, it is unlikely that seminal studies were excluded for several reasons: i) a comprehensive search strategy was developed by an international, interprofessional team with relevant methodological and clinical expertise with the assistance of an experienced medical sciences librarian; ii) the search strategy was piloted and refined prior to being used; and iii) the review was conducted in a systematic fashion (i.e. using groups of two independent reviewers and data extractors). While it was intended that only original quantitative research data from studies utilizing SMob would be reported, it was not always clear as to whether reported data were previously published in part or fully. However, in instances where this was unclear, the decision was made to include the data. This decision ensured that the current review reported exhaustively on all studies reporting biomechanical parameters of SMob. It is recommended that secondary analyses of data are transparently reported as such (57,58), with citation of the original publication, allowing readers to identify that the data has been previously published and to interpret for themselves the impact of the re-reported data.

## CONCLUSION

This study has, as a first step, synthesised the current state of manually applied SMob biomechanical parameter reporting. Most studies reported on SMob delivered to human lumbar or cervical spines, with peak force the most commonly reported parameter. Other reported parameters included duration, frequency and force amplitude. These findings highlight that considerable variability exists in the literature regarding SMob biomechanical parameters. Future studies should focus on the detailed reporting of biomechanical parameters which may facilitate the systematic investigation of dose-response effects clinically and the future development of reference standards (e.g. ranges of forces) for optimal intervention delivery.

## Data Availability

The datasets used and/or analysed during the current study are available from the corresponding author on reasonable request.

## ACKNOWLEDGEMENTS

The authors would like to acknowledge Dr. XX (The University of XX) for her assistance with the literature search.

## Notes

### Competing Interest Statement

The authors have declared no competing interest.

### Clinical Trial

N/A.

### Funding Statement

The author(s) received no specific funding for this work.

### Author Declarations

Ethics approval was not required for this scoping literature review.

## REFERENCES

1. Hoy DG, Protani M, De R, Buchbinder R. The epidemiology of neck pain. Best Pract Res Clin Rheumatol. 2010 Dec;24(6):783–92.

2. Hoy D, Bain C, Williams G, March L, Brooks P, Blyth F, et al. A systematic review of the global prevalence of low back pain. Arthritis Rheum. 2012 Jun;64(6):2028–37.

3. Meucci RD, Fassa AG, Faria NMX. Prevalence of chronic low back pain: systematic review. Rev Saúde Pública. 2015;49.

4. Kazeminasab S, Nejadghaderi SA, Amiri P, Pourfathi H, Araj-Khodaei M, Sullman MJM, et al. Neck pain: global epidemiology, trends and risk factors. BMC Musculoskelet Disord. 2022 Jan 3;23(1):26.

5. Safiri S, Kolahi AA, Cross M, Hill C, Smith E, Carson-Chahhoud K, et al. Prevalence, Deaths, and Disability-Adjusted Life Years Due to Musculoskeletal Disorders for 195 Countries and Territories 1990–2017. Arthritis Rheumatol. 2021 Apr 1;73(4):702–14.

6. Froud R, Patterson S, Eldridge S, Seale C, Pincus T, Rajendran D, et al. A systematic review and meta-synthesis of the impact of low back pain on people’s lives. BMC Musculoskelet Disord. 2014 Feb 21;15(1):50.

7. Safiri S, Kolahi AA, Hoy D, Buchbinder R, Mansournia MA, Bettampadi D, et al. Global, regional, and national burden of neck pain in the general population, 1990-2017: systematic analysis of the Global Burden of Disease Study 2017. BMJ. 2020 Mar 26;368:m791.

8. Vos T, Lim SS, Abbafati C, Abbas KM, Abbasi M, Abbasifard M, et al. Global burden of 369 diseases and injuries in 204 countries and territories, 1990–2019: a systematic analysis for the Global Burden of Disease Study 2019. The Lancet. 2020 Oct 17;396(10258):1204–22.

9. Dieleman JL, Cao J, Chapin A, Chen C, Li Z, Liu A, et al. US Health Care Spending by Payer and Health Condition, 1996-2016. JAMA. 2020 Mar 3;323(9):863–84.

10. Chen N, Fong DYT, Wong JYH. Health and Economic Outcomes Associated With Musculoskeletal Disorders Attributable to High Body Mass Index in 192 Countries and Territories in 2019. JAMA Netw Open. 2023 Jan 20;6(1):e2250674–e2250674.

11. Power JD, Perruccio AV, Paterson JM, Canizares M, Veillette C, Coyte PC, et al. Healthcare Utilization and Costs for Musculoskeletal Disorders in Ontario, Canada. J Rheumatol. 2022 Jul;49(7):740–7.

12. Corp N, Mansell G, Stynes S, Wynne-Jones G, Morsø L, Hill JC, et al. Evidence-based treatment recommendations for neck and low back pain across Europe: A systematic review of guidelines. Eur J Pain. 2021 Feb 1;25(2):275–95.

13. Risetti M, Gambugini R, Testa M, Battista S. Management of non-specific thoracic spine pain: a cross-sectional study among physiotherapists. BMC Musculoskelet Disord. 2023 May 19;24(1):398.

14. Beliveau PJH, Wong JJ, Sutton DA, Simon NB, Bussières AE, Mior SA, et al. The chiropractic profession: a scoping review of utilization rates, reasons for seeking care, patient profiles, and care provided. Chiropr Man Ther. 2017 Nov 22;25(35).

15. Babatunde OO, Jordan JL, Van der Windt DA, Hill JC, Foster NE, Protheroe J. Effective treatment options for musculoskeletal pain in primary care: A systematic overview of current evidence. PloS One. 2017;12(6):e0178621.

16. Lin I, Wiles L, Waller R, Goucke R, Nagree Y, Gibberd M, et al. What does best practice care for musculoskeletal pain look like? Eleven consistent recommendations from high-quality clinical practice guidelines: systematic review. Br J Sports Med. 2020 Jan 1;54(2):79.

17. Foster NE, Hartvigsen J, Croft PR. Taking responsibility for the early assessment and treatment of patients with musculoskeletal pain: a review and critical analysis. Arthritis Res Ther. 2012 Feb 29;14(1):205.

18. George SZ, Fritz JM, Silfies SP, Schneider MJ, Beneciuk JM, Lentz TA, et al. Interventions for the Management of Acute and Chronic Low Back Pain: Revision 2021. J Orthop Sports Phys Ther. 2021 Nov 1;51(11):CPG1–60.

19. Blanpied PR, Gross AR, Elliott JM, Devaney LL, Clewley D, Walton DM, et al. Neck Pain: Revision 2017. J Orthop Sports Phys Ther. 2017 Jul 1;47(7):A1–83.

20. Bussieres AE, Stewart G, Al-Zoubi F, Decina P, Descarreaux M, Haskett D, et al. Spinal Manipulative Therapy and Other Conservative Treatments for Low Back Pain: A Guideline From the Canadian Chiropractic Guideline Initiative. J Manip Physiol Ther. 2018 Mar 29;

21. Bussieres AE, Stewart G, Al-Zoubi F, Decina P, Descarreaux M, Hayden J, et al. The treatment of neck pain-associated disorders and whiplash-associated disorders: a clinical practice guideline. J Manipulative Physiol Ther. 2016 Oct;39(8):523–64.

22. Snodgrass SJ, Rivett DA, Robertson VJ. Manual Forces Applied During Posterior-to-Anterior Spinal Mobilization: A Review of the Evidence. J Manipulative Physiol Ther. 2006 May;29(4):316–29.

23. Maitland, GD. Vertebral manipulation. 5th ed. London: Butterworth; 1986.

24. Maitland, GD. Maitland’s vertebral manipulation. 6th ed. Oxford, England: Butterworth-Heinemann; 2001.

25. Hegedus EJ, Goode A, Butler RJ, Slaven E. The neurophysiological effects of a single session of spinal joint mobilization: does the effect last? J Man Manip Ther. 2011 Aug;19(3):143–51.

26. Sterling M, Jull G, Wright A. Cervical mobilisation: concurrent effects on pain, sympathetic nervous system activity and motor activity. Man Ther. 2001 May;6(2):72–81.

27. Ali MN, Sethi K, Noohu MM. Comparison of two mobilization techniques in management of chronic non-specific low back pain. J Bodyw Mov Ther. 2019 Oct;23(4):918–23.

28. Schmid A, Brunner F, Wright A, Bachmann LM. Paradigm shift in manual therapy? Evidence for a central nervous system component in the response to passive cervical joint mobilisation. Man Ther. 2008 Oct;13(5):387–96.

29. Bialosky JE, Bishop MD, Price DD, Robinson ME, George SZ. The mechanisms of manual therapy in the treatment of musculoskeletal pain: a comprehensive model. Man Ther. 2009;14(5):531–8.

30. Alonso-Perez JL, Lopez-Lopez A, La Touche R, Lerma-Lara S, Suarez E, Rojas J, et al. Hypoalgesic effects of three different manual therapy techniques on cervical spine and psychological interaction: A randomized clinical trial. J Bodyw Mov Ther. 2017;21(4):798–803.

31. Hidalgo B, Pitance L, Hall T, Detrembleur C, Nielens H. Short-term effects of Mulligan mobilization with movement on pain, disability, and kinematic spinal movements in patients with nonspecific low back pain: a randomized placebo-controlled trial. J Manipulative Physiol Ther. 2015 Aug;38(6):365–74.

32. Pasquier M, Daneau C, Marchand AA, Lardon A, Descarreaux M. Spinal manipulation frequency and dosage effects on clinical and physiological outcomes: a scoping review. Chiropr Man Ther. 2019 May 22;27(1):23.

33. Matesanz-García L, Schmid AB, Cáceres-Pajuelo JE, Cuenca-Martínez F, Arribas-Romano A, González-Zamorano Y, et al. Effect of Physiotherapeutic Interventions on Biomarkers of Neuropathic Pain: A Systematic Review of Preclinical Literature. J Pain. 2022 Nov;23(11):1833– 55.

34. Lascurain-Aguirrebeña I, Newham D, Critchley DJ. Mechanism of Action of Spinal Mobilizations: A Systematic Review. Spine. 2016 Jan;41(2):159–72.

35. Aoyagi K, Heller D, Hazlewood D, Sharma N, dos Santos M. Is spinal mobilization effective for low back pain?: A systematic review. Complement Ther Clin Pract. 2019;34:51–63.

36. Bjornsdottir SV, Kumar S. Posteroanterior spinal mobilization: state of the art review and discussion. Disabil Rehabil. 1997 Feb;19(2):39–46.

37. Tricco AC, Lillie E, Zarin W, O’Brien KK, Colquhoun H, Levac D, et al. PRISMA Extension for Scoping Reviews (PRISMA-ScR): Checklist and Explanation. Ann Intern Med. 2018 Oct 2;169(7):467–73.

38. Cooke A, Smith D, Booth A. Beyond PICO: the SPIDER tool for qualitative evidence synthesis. Qual Health Res. 2012 Oct;22(10):1435–43.

39. Ouzzani M, Hammady H, Fedorowicz Z, Elmagarmid A. Rayyan-a web and mobile app for systematic reviews. Syst Rev. 2016 Dec 5;5(1):210.

40. Mercier MA, Rousseau P, Funabashi M, Descarreaux M, Pagé I. Devices Used to Measure Force-Time Characteristics of Spinal Manipulations and Mobilizations: A Mixed-Methods Scoping Review on Metrologic Properties and Factors Influencing Use. Front Pain Res [Internet]. 2021;2. Available from: https://www.frontiersin.org/articles/10.3389/fpain.2021.755877

41. Hoffmann TC, Glasziou PP, Boutron I, Milne R, Perera R, Moher D, et al. Better reporting of interventions: template for intervention description and replication (TIDieR) checklist and guide. BMJ. 2014 Mar 7;348:g1687.

42. Mikhail J, Funabashi M, Descarreaux M, Page I. Assessing forces during spinal manipulation and mobilization: factors influencing the difference between forces at the patient-table and clinician-patient interfaces. Chiropr Man Ther. 11 10;28(1):57.

43. Kirstukas SJ, Backman JA. Physician-applied contact pressure and table force response during unilateral thoracic manipulation. J Manipulative Physiol Ther. 1999 Jun;22(5):269–79.

44. Passmore SR, Malone Q, MacNeil B, Sanli E, Gonzalez D. Differing Characteristics of Human-Shaped Visual Stimuli Affect Clinicians’ Dosage of a Spinal Manipulative Thrust on a Low-Fidelity Model: A Cross-Sectional Study. J Manipulative Physiol Ther. 2022 Mar;45(3):171–8.

45. Snodgrass SJ, Rivett DA, Robertson VJ, Stojanovski E. A comparison of cervical spine mobilization forces applied by experienced and novice physiotherapists. J Orthop Sports Phys Ther. 2010 Jul;40(7):392–401.

46. Harms MC, Innes SM, Bader DL. Forces measured during spinal manipulative procedures in two age groups. Rheumatology. 1999 Mar;38(3):267–74.

47. Chiradejnant A, Latimer J, Maher CG. Forces applied during manual therapy to patients with low back pain. J Manipulative Physiol Ther. 2002 Jul;25(6):362–9.

48. Kearns GA, Brismée JM, Riley SP, Wang-Price S, Denninger T, Vugrin M. Lack of standardization in dry needling dosage and adverse event documentation limits outcome and safety reports: a scoping review of randomized clinical trials. J Man Manip Ther. 2023 Apr;31(2):72–83.

49. Groeneweg R, Rubinstein SM, Oostendorp RAB, Ostelo RWJG, van Tulder MW. Guideline for Reporting Interventions on Spinal Manipulative Therapy: Consensus on Interventions Reporting Criteria List for Spinal Manipulative Therapy (CIRCLe SMT). J Manipulative Physiol Ther. 2017;40(2):61–70.

50. Alvarez G, Sola I, Sitja-Rabert M, Fort-Vanmeerhaeghe A, Gich I, Fernandez C, et al. A methodological review revealed that reporting of trials in manual therapy has not improved over time. J Clin Epidemiol. 5;121:32–44.

51. Descarreaux M, Dugas C, Raymond J, Normand MC. Kinetic analysis of expertise in spinal manipulative therapy using an instrumented manikin. J Chiropr Med. 2005 Mar 1;4(2):53–60.

52. Gorgos KS, Wasylyk NT, Van Lunen BL, Hoch MC. Inter-clinician and intra-clinician reliability of force application during joint mobilization: a systematic review. Man Ther. 2014 Apr;19(2):90– 6.

53. Coulter ID, Crawford C, Vernon H, Hurwitz EL, Khorsan R, Booth MS, et al. Manipulation and Mobilization for Treating Chronic Nonspecific Neck Pain: A Systematic Review and Meta-Analysis for an Appropriateness Panel. Pain Physician. 2019 Mar;22(2):E55–70.

54. Coulter ID, Crawford C, Hurwitz EL, Vernon H, Khorsan R, Suttorp Booth M, et al. Manipulation and mobilization for treating chronic low back pain: a systematic review and meta-analysis. Spine J Off J North Am Spine Soc. 5;18(5):866–79.

55. Snodgrass SJ, Rivett DA, Sterling M, Vicenzino B. Dose optimization for spinal treatment effectiveness: a randomized controlled trial investigating the effects of high and low mobilization forces in patients with neck pain. J Orthop Sports Phys Ther. 2014 Mar;44(3):141–52.

56. Gorrell LM, Beath K, Engel RM. Manual and instrument applied cervical manipulation for mechanical neck pain: a randomized controlled trial. J Manipulative Physiol Ther. 2016;39(5):319– 29.

57. Swart E, Schmitt J. STandardized Reporting Of Secondary data Analyses (STROSA) - Vorschlag für ein Berichtsformat für Sekundärdatenanalysen. Qualitätsmessung. 2014 Jan 1;108(8):511–6.

58. Schulz KF, Altman DG, Moher D, the CONSORT Group. CONSORT 2010 Statement: updated guidelines for reporting parallel group randomised trials. BMC Med. 2010 Mar 24;8(1):18.

